# A Performance Evaluation of Computerised Antepartum Fetal Heart Rate Monitoring: The Dawes-Redman Algorithm at Term

**DOI:** 10.1101/2024.02.12.24302705

**Authors:** Gabriel Davis Jones, Beth Albert, William Cooke, Manu Vatish

**Author notes:** Corresponding author: Gabriel Davis Jones.

## Abstract

**Objectives:** This study aims to rigorously evaluate the Dawes-Redman computerised cardiotocography algorithm’s effectiveness in assessing antepartum fetal wellbeing. It focuses on analysing the algorithm’s performance using extensive clinical data, examining accuracy, sensitivity, specificity, and predictive values in various scenarios. The objectives include assessing the algorithm’s reliability in identifying fetal wellbeing across different risk prevalences, its efficacy in the context of temporal proximity to delivery, and its performance across ten specific adverse pregnancy outcomes. This comprehensive evaluation seeks to clarify the algorithm’s utility and limitations in contemporary obstetric practice, particularly in high-risk pregnancy scenarios.

**Methods:** Antepartum fetal heart rate recordings from term singleton pregnancies between 37 and 42 gestational weeks were extracted from the Oxford University Hospitals database, spanning 1991 to 2021. Traces with significant data gaps or incomplete Dawes-Redman analyses were excluded. For the ten adverse outcomes, only traces performed within 48 hours prior to delivery were considered, aligning with clinical decision-making practices. A healthy cohort was established using rigorous inclusion and exclusion criteria based on clinical indicators. Propensity score matching, controlling for gestational age and fetal sex, ensured balanced comparisons between healthy and adverse outcome cohorts. The Dawes-Redman algorithm’s categorisation of FHR traces as either ‘criteria met’ (an indicator of wellbeing) or ‘criteria not met’ (indicating a need for further evaluation) informed the evaluation of predictive performance metrics. Performance was assessed using accuracy, sensitivity, specificity, and predictive values (PPV, NPV), adjusted for various risk prevalences.

**Results:** 4,196 term antepartum FHR traces were identified, matched by fetal sex and gestational age. The Dawes-Redman algorithm showed a high sensitivity of 91.7% for detecting fetal wellbeing. However, specificity for adverse outcomes was low at 15.6%. The PPV varied with population prevalence, high in very low-risk settings (99.1%) and declined with increased risk. Temporal proximity to delivery indicated robust sensitivity (>91.0%). Specificity notably decreased over time, impacting the algorithm’s discriminative power for identifying adverse outcomes. Across different adverse conditions, the algorithm’s performance remained consistent, with high sensitivity but varying NPVs, confirming its utility in detecting fetal wellbeing rather than adverse outcomes.

**Conclusion:** These findings reveal the Dawes-Redman algorithm is effective for detecting fetal wellbeing in term pregnancies, evidenced by its high sensitivity and PPV. However, its low specificity suggests limitations in its ability to identify fetuses at risk of adverse outcomes. The predictive accuracy of the algorithm is significantly affected by the prevalence of healthy pregnancies within the population. Clinical interpretation of FHR traces that do not satisfy the Dawes-Redman criteria should be approached with caution, as they do not necessarily correlate with heightened risk. While the algorithm proves reliable for its primary objective in low-risk contexts, the development of algorithms optimised for high-risk pregnancy scenarios remains an area for future enhancement.

## Introduction

Fetal heart rate (FHR) monitoring (the ‘non-stress test’ or ‘cardiotocography’) is one of the commonest obstetric investigations performed worldwide.^1^ It involves the non-invasive application of an ultrasound transducer to the maternal abdomen to continuously evaluate the FHR, enabling real-time, continuous assessment of fetal physiology. Patterns within the FHR trace are associated with the central and peripheral nervous systems and fetal endocrine activity^2,3^. These patterns are used to evaluate fetal brain health and overall wellbeing.^4,5^ FHR monitoring in the third trimester before labour (the antepartum) is frequently performed to assess whether a baby is at risk of an adverse outcome or death, indicating early pregnancy intervention is required.^6,7^

Antepartum FHR monitoring has been a mainstay of pregnancy care since the 1960s. Traces are traditionally interpreted visually, however the reproducibility and reliability of human visual analysis has remained consistently poor.^8^ Expert clinical evaluation fails to accurately identify between 35–92% of fetal heart rate patterns.^9,10^ Inter- and intra-observer agreement between experts has been estimated as low as 29% while false positive rates for identifying an at-risk fetus are as high as 60%.^11-16^ Human misinterpretation of these patterns has been directly implicated in avoidable early pregnancy intervention, increased adverse pregnancy outcomes (including fetal death) and is a major source of medicolegal litigation globally.^17-20^ Efforts to standardize visual evaluation methods in antepartum fetal heart rate monitoring have faced issues with performance, reproducibility and clinician consensus.^20-24^

Professors Geoffrey Dawes and Christopher Redman at the University of Oxford began developing a computerised algorithm in the 1978 to address this.^25,26^ The goal was to use automated FHR analysis to identify fetuses in a state of wellbeing. The algorithm identifies a baseline FHR within the trace which it then uses to extract several clinically-validated patterns: the basal FHR, accelerations, decelerations, episodes of high or low variation, short-term variability (STV) and long-term variability (LTV). These patterns have been associated with healthy and/or adverse fetal outcomes.^27^ Numerical analysis of these patterns was then used to develop a set of ten criteria for fetal wellbeing: the Dawes-Redman Criteria.^28^ If all criteria are fulfilled, the trace is classified as ‘criteria met’, the fetus deemed ‘healthy’ and the operator advised to cease FHR monitoring. If the trace fails to meet all ten criteria after 60 minutes, it is categorised as ‘criteria not met’, analysis halts and guidance is given to seek expert clinical opinion.

The first commercial version of the Dawes-Redman algorithm was released in 1989 and has since become ubiquitous in antepartum FHR monitoring globally.^28^ Following publication of the algorithm in 2002, manufacturers of FHR monitors have incorporated the algorithm and derivatives thereof into their systems.^28-31^ In the UK, it is currently found in over 120 hospitals while internationally it is sold in more than 110 countries. It is incorporated into local and national guidelines.^32,33^ Clinical decisions regarding the pregnancy are made using its output. While the algorithm was designed to identify fetuses in a state of wellbeing, its results have also been used to identify at-risk pregnancies.^34,35^ Several studies have evaluated components of the Dawes-Redman algorithm (e.g. STV or LTV) in high-risk pregnancies (e.g. hypoxaemia^36^, acidaemia^37^, intrauterine or neonatal death^38^). However, the algorithm was not designed for this purpose. There has never been a large-scale, robust analysis of the algorithm’s performance at its actual objective, despite its global prominence in obstetric care.^32^

We have undertaken the first evaluation of the Dawes-Redman algorithm for its intended and most common purpose at term: identifying antepartum fetuses in a state of wellbeing. We utilised high-fidelity clinical data from the last 30 years of Dawes-Redman use at the John Radcliffe Hospital in Oxford, UK. We developed two cohorts of antepartum FHR traces acquired at term: those from pregnancies with healthy fetal outcomes and another with high-risk adverse outcomes. Performance was evaluated with six metrics important in clinical practice: accuracy, sensitivity, specificity, positive predictive value, negative predictive value and the F1 score. We have evaluated the general performance of the algorithm between 0–48 hours prior to delivery and whether temporal proximity to delivery and discrete adverse outcomes affect the reliability of the Dawes-Redman algorithm.

## Methods

### Data processing, cohort development and extraction of fetal heart rate patterns

We extracted raw digital antepartum fetal heart rate (FHR) traces from the Oxford University Hospitals maternity database at the John Radcliffe Hospital (Oxford, United Kingdom) between the 1st of January 1991 and 31^st^ of December 2021. Traces were acquired from singleton pregnancies between 37^+0^ and 41^+6^ gestational weeks for which associated clinical outcome information for the mother and baby were available. This study was approved by the Ethics Committee in Joint Research Office, Research and Development Department, Oxford University Hospitals NHS Trust: 13/SC/0153. Each trace had previously undergone Dawes-Redman analysis as part of standard clinical practice. Traces missing >30% of their signal information or had their Dawes-Redman analysis aborted before evaluation could complete were removed.

We identified ten adverse pregnancy outcomes in the database. These included acidaemia, asphyxia, birthweight <3^rd^ centile for gestational age, extended special care (SCBU) admission, hypoxic ischaemic encephalopathy, low Apgar scores, neonatal sepsis, perinatal infections, respiratory conditions and antepartum or intrapartum stillbirth. Outcome definitions are provided in Supplementary Table 4. In cases of adverse pregnancy outcomes, only FHR traces acquired within 48 hours prior to delivery of the fetus were selected. Antepartum FHR monitoring is performed for myriad indications. Thus, it is not accurate to presume monitoring performed over the course of a pregnancy pertains to a consistent clinical indication or presentation. Selecting for traces collected well before an adverse outcome occurs, without corroborating clinical evidence, would imply all traces were conducted in the presence of (the same) pathology. Constraining the timeframe to within 48 hours before delivery aids in mitigating this assumption. This approach also aligns more closely with clinical practice whereby the outcome of FHR monitoring is used to inform more immediate decision making.

To establish a cohort of healthy outcome pregnancies for comparison, a set of inclusion and exclusion criteria were utilised. Pregnancies with antepartum FHR traces were included in the healthy outcome pregnancy cohort if they met the following criteria: liveborn singleton baby, gestational age at delivery between 37^+0^ and 41^+6^ weeks, maternal age at booking between 18 to 39 years, birthweight ≥10^th^ centile, duration of labour less than 24 hours, an Apgar Score of ≥4 at 1 minute and ≥ 7 at 5 minutes and umbilical venous ±arterial pH within the normal range (arterial pH >7.13 and base deficit <10.0 for babies delivered via caesarean section without labour; arterial pH >7.05 and base deficit <14.0 for babies who experienced labour), and a cerebroplacental ratio greater than 1.5 at 36 weeks.^39^ The latter two criteria only applied if the corresponding data were available.

Pregnancies with antepartum FHR traces were excluded from the healthy outcome cohort for any of the following reasons: neonatal death within 3 months following delivery, emergency caesarean section delivery, breech presentation, any requirement for neonatal resuscitation, SCBU admission following delivery or neonatal cooling, hypertensive disorders of pregnancy such as pre-eclampsia and clinically-suspected but unconfirmed infection or sepsis.

Antepartum FHR traces from the healthy outcome cohort were then matched with corresponding traces from pregnancies with confirmed adverse outcomes. One-to-one matching was performed using propensity score matching controlling for the gestational age when FHR monitoring was performed and fetal sex.^40^ This ensures each case in the healthy outcome cohort has a directly comparable counterpart in the adverse outcome cohort, thereby minimising the potential for bias in the comparison. One-to-one matching also facilitates modelling of outcome prevalences by creating balanced groups, which enables more accurate estimations of the effect size attributable to the health outcomes being studied. Controlling for gestational age is vital, as it directly influences FHR characteristics. Variations in FHR patterns can be attributed to the developmental stage of the fetus.^41-43^ Ignoring this variable could introduce bias and confound results. Similarly, fetal sex is to control for, given male and female fetuses exhibit different physiological responses, intrauterine stress.^44,45^

To ensure precision in matching, a maximum difference of 0.05 in propensity scores was allowed between matched pairs to constrain the similarity of their characteristics. Each trace from a pregnancy was used only once to avoid duplication. Following matching, the average differences between the gestational ages at monitoring and fetal sexes for between the cohorts were calculated to confirm the adequacy of the matching process.

### Performance analysis

We categorised Dawes-Redman algorithm results as ‘positive’ when the algorithm assigned the FHR trace as ‘criteria met’ (e.g., the fetus is in a state of wellbeing) and negative when designated as ‘criteria not met’. This terminology stems from the algorithm’s foundational purpose for identifying ‘normal’ FHR traces. Accordingly, we label a pregnancy as ‘positive’ if it aligned with the healthy outcome cohort and ‘negative’ otherwise. True positives therefore refer to cases where both the FHR trace and pregnancy status are classified as positive. False positives are when the trace is deemed positive despite the pregnancy belonging to the adverse outcome cohort. True negatives occur when the algorithm identifies the trace as negative in adverse outcome pregnancies, while false negatives represent instances where the trace is associated with the healthy outcome cohort yet the algorithm classified the trace as ‘criteria not met’, i.e. negative. A tabular representation of these terms is presented in Supplementary Table 5.

Accuracy, sensitivity, specificity, positive predictive value (PPV), negative predictive value (NPV) and the F1 score were calculated for each analysis. Accuracy represents the proportion of total classifications (both positive and negative) that are correct. In this context, it evaluates how accurately the algorithm classifies FHR traces and aligns them with the corresponding pregnancy outcome. Sensitivity reflects the ability of the algorithm to correctly identify ‘positive’ cases, i.e., instances where both the FHR trace is classified as ‘criteria met’ and the pregnancy outcome is healthy. Specificity assesses the system’s capability to correctly identify ‘negative’ cases, where both the FHR trace is classified as ‘criteria not met’ and the pregnancy belongs to the adverse outcome cohort. PPV indicates the probability that a ‘positive’ result (FHR trace classified as criteria met) corresponds to a healthy pregnancy. NPV denotes the probability that a ‘negative’ result (FHR trace classified as ‘criteria not met’) corresponds to an adverse pregnancy outcome. The F1 score is the harmonic mean of sensitivity and PPV, enabling a more balanced assessment of the system’s ability to identify healthy pregnancies.

Accurate appraisal of the PPV and NPV is dependent on the underlying prevalence of the outcome of interest within a given population, here healthy pregnancies.^46^ For clinical contextualisation, we evaluated the PPV and NPV across several theoretical risk strata: a normality prevalence of 99% (very low risk), 90% (low risk), 80% (medium risk) and 70% (high risk). The enables adjustment of the PPV and NPV based on the specified prevalence, facilitating contextualisation of performance with scenarios more aligned with obstetric practice.

To circumvent potential bias stemming from pregnancies with multiple FHR traces within a particular data group, lower numbers of traces at different gestational ages or imbalance in specific adverse outcomes, stratified bootstrapping was employed for each analysis.^47^ A balanced, random sample consisting of normal healthy pregnancies and adverse outcome pregnancies was drawn from each gestational age and time before delivery (in the case of adverse pregnancy outcomes), ensuring a fair representation. Performance metrics were then computed for each random sample. This was repeated 100,000 times with the mean and 95% confidence intervals derived from these values for each performance metric.

Three analyses were conducted. We first evaluated the performance of the Dawes-Redman algorithm for all term antepartum FHR traces acquired between 0–48 hours prior to delivery. We then evaluated whether temporal proximity to delivery affected performance. We split the data into two time windows: cases in which the adverse outcome antepartum FHR trace was acquired between 0 and 23 hours and 59 minutes prior to delivery (0–24 hours) and those in which the adverse outcome trace was acquired between 24 and 47 hours and 59 minutes (24–48 hours) prior to delivery. We then compared the performance in both windows. Finally, we investigated if the ability of the Dawes-Redman algorithm to identify fetuses in a state of wellbeing differed meaningfully by adverse outcome. We calculated the performance metrics for each discrete outcome comprising the adverse outcome cohort separately for all traces between 0–48 hours.

This methodological framework was devised to not only scrutinise the data from multiple perspectives – general performance, temporal proximity to delivery and adverse outcomes – but also to ensure a balanced and unbiased representation of all pregnancies, thereby aiming to furnish a robust and insightful analysis of the Dawes-Redman algorithm’s performance in clinical practice.

### Statistical analysis

Discrete variables are presented as numbers (with interquartile ranges) and percentages while continuous variables are listed as mean and 95% confidence intervals (95% CI). Performance metrics are expressed in percentages. Categorical variables were compared using the Chi-square test while continuous variables were compared using the Mann-Whitney U test with a significant threshold of 0.05. Sensitivity, specificity, PPV, NPV and the F1 score were calculated using standard methods.^48^ PPV, NPV and their confidence intervals were calculated using the method described by Mercaldo *et al*.^49^ Cohen’s f was employed to determine effect sizes, with f values of 0.10 or less denoting a small effect, values up to 0.25 indicating a medium effect and values up to 0.40 representing a large effect.^50^ Analysis was performed using Python (version 3.9.17) with the Pandas (version 1.5.3), NumPy (version 1.23.5), Matplotlib (version 3.7.1) and SciPy (version 1.10.1) packages.

## Results

A total of 4,196 antepartum FHR traces from 1,820 healthy outcome and 1,560 adverse outcome pregnancies recorded at term were used for analysis. 1:1 matching by fetal sex and gestational age at recording resulted in 2,098 traces from healthy outcome pregnancies and 2,098 traces from adverse outcome pregnancies. 49.7% (1,042) of the traces were from adverse outcome pregnancies delivered within 24 hours after the trace was performed. 51.3% (1,056) of the traces were from adverse outcome pregnancies delivered within 24–48 hours. Supplementary Tables 1 and 2 show the baseline characteristics of infants and mothers. Supplementary Table 3 describes the frequency of each adverse outcome.

Maternal age was comparable, with a median of 31.0 years in both groups (p=0.39). Healthy outcome pregnancies reported a median maternal BMI of 23.3 kg/m^2^ and adverse outcomes a median BMI of 25.3kg/m^2^ (p<0.01). There was negligible variance in both viable and non-viable parity, with medians of 1.0 and 0.0 respectively in both groups (p-value <0.01, effect size 0.1). Maternal smoking status at delivery differed. The healthy outcomes group had no current smokers, more ex-smokers (18.4% vs. 9.1%), and never-smokers (74.1% vs. 39.4%) than the adverse outcomes group. The mode of labour onset showed significant differences. Inductions were more common in adverse outcome pregnancies (800, 51.3%) compared to healthy outcomes (613, 33.7%, p<0.01). Adverse outcome pregnancies did not experience labour more frequently (366, 23.5%) compared to healthy outcome pregnancies (231, 12.7%, p<0.01). Spontaneous labour was reported in 976 (53.6%) of healthy outcome pregnancies against 394 (25.3%, p<0.01) of adverse outcome pregnancies.

Male and female births were proportionally similar across healthy and adverse outcome pregnancies (p=0.46). Birthweight was significantly lower in the adverse outcome group (p<0.001). The median gestational age at monitoring was comparable between groups (p=0.99), however gestational age at delivery was earlier in adverse outcome pregnancies (39^+3^ vs 40^+1^, p<0.001). Apgar scores at 1 minute were lower in the adverse outcome pregnancies, with diminished but still significant differences at 5 and 10 minutes (p<0.001). Elective caesareans were more common in adverse outcome pregnancies (p<0.001) while spontaneous vertex delivery was more common in healthy outcome pregnancies (p<0.001).

### Performance evaluation

Analysis of the balanced cohorts revealed that 45.9% (95% CI: 45.2–46.5%) of the traces meeting the Dawes-Redman criteria were associated with healthy pregnancy outcomes (true positives), while 42.2% (95% CI: 41.4–43.0%) corresponded to adverse pregnancy outcomes (false positives). In cases where the Dawes-Redman criteria were not met, 4.1% (95% CI: 3.5–4.8%) were associated with healthy outcomes (false negatives), and 7.8% (95% CI: 7.0–8.6%) to adverse outcomes (true negatives). Table 1 shows the confusion matrix.

**Table 1:**
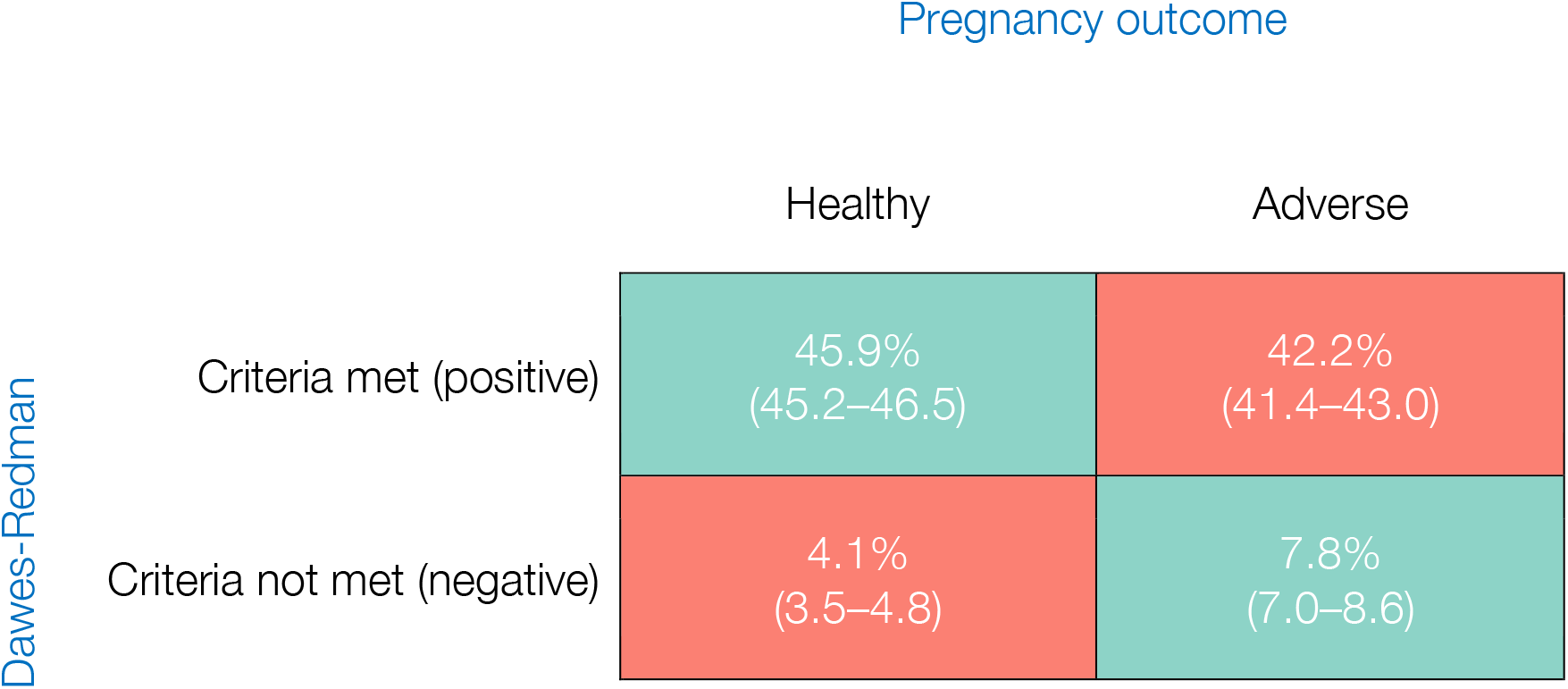
Confusion Matrix for Dawes-Redman Analysis at term. 4,196 FHR traces from healthy (N=1,820) and adverse (N=1,560) outcome pregnancies were analysed. The datasets were balanced for fetal sex and gestational age when the trace was performed, resulting in a balanced dataset of 2,098 traces in each group. For the adverse pregnancy outcome group, only traces acquired within 48 hours prior to delivery were used. See Supplementary Table 2 for a definition of the confusion matrix. Values are expressed as a percent (%) with 95% confidence intervals.

The overall accuracy of the Dawes-Redman algorithm was 53.7% (95% CI: 52.6–54.7), which reflects the proportion of total cases correctly identified as either normal or adverse outcome. Sensitivity, in this case, is the algorithm’s ability to correctly identify healthy outcome pregnancies, which was 91.7% (95% CI: 90.4–92.9). This high sensitivity indicates the algorithm is effective at detecting pregnancies where the fetus is in a state of wellbeing. However, the specificity, here the ability to correctly identify pregnancies with adverse outcomes, is considerably lower at 15.6% (95% CI: 14.0–17.3), suggesting the algorithm has limited effectiveness in correctly identifying cases where the fetus is not in a state of wellbeing (Table 2).

**Table 2:** Performance of the Dawes-Redman algorithm at identifying normal healthy pregnancies and adverse outcome pregnancies at term. Each adverse outcome FHR trace was from a pregnancy that was subsequently delivered within 48 hours of monitoring. Each was matched by gestational age and fetal sex with a corresponding trace from a healthy outcome pregnancy, not necessarily delivered within the same time window. Four theoretical risk groups were evaluated: very low risk, low risk, medium risk and high risk to demonstrate how prevalence of outcomes affects performance. These groups denote the underlying prevalence of a ‘normal’ and ‘adverse outcome’ pregnancy and demonstrate how the performance of the algorithm changes when the prevalence of normality decreases in the population while the prevalence of adverse outcomes increase. As prevalence of normal pregnancies decrease, the performance of the algorithm declines substantially from an F1 score of 95.2% in the very low risk group to 80.5% in the high risk group.

| Performance Metric | % (95% CI) |
| --- | --- |
| Accuracy | 53.7 (52.6–54.7) |
| Sensitivity | 91.7 (90.4–92.9) |
| Specificity | 15.6 (14.0–17.3) |
| **Very low risk** 99% normal outcomes, 1% adverse outcomes |  |
| Positive predictive value | 99.1 (99.1–99.1) |
| Negative predictive value | 1.9 (1.6–2.3) |
| F1 Score | 95.2 (94.5–95.9) |
| **Low risk** 90% normal outcomes, 10% adverse outcomes |  |
| Positive predictive value | 90.7 (90.5–90.9) |
| Negative predictive value | 17.4 (14.8–20.2) |
| F1 Score | 91.2 (90.5–91.9) |
| **Medium risk** 80% normal outcomes, 20% adverse outcomes |  |
| Positive predictive value | 81.3 (80.9–81.7) |
| Negative predictive value | 32.1 (28.2–36.3) |
| F1 Score | 86.2 (85.5–86.9) |
| **High risk** 70% normal outcomes, 30% adverse outcomes |  |
| Positive predictive value | 71.7 (71.2–72.2) |
| Negative predictive value | 44.7 (40.2–49.5) |
| F1 Score | 80.5 (79.8–81.2) |

The Dawes-Redman algorithm’s efficacy in predicting healthy pregnancy outcomes exhibited a decline with decreasing population prevalence of healthy pregnancies. Within the very low-risk cohort, where the prevalence of healthy outcomes was 99%, the algorithm demonstrated a high positive predictive value (PPV) of 99.1% (95% CI: 99.1– 99.1). However, its negative predictive value (NPV) was markedly low at 1.9% (95% CI: 1.6–2.3) and the F1 Score, the harmonic mean of sensitivity and PPV, was 95.2% (95% CI: 94.5–95.9). As risk stratification increased, with the proportion of healthy pregnancies decreasing from 90% to 70% respectively, there was a proportional decline in PPV, from 90.7% to 71.7% and an increase in NPV from 17.4% to 44.7%. This trend was accompanied by a corresponding decrease in the F1 Score from 91.2% to 80.5% (Table 2). We demonstrate the principle that PPV and NPV are directly associated with population prevalence in Figure 1.

**Figure 1:**
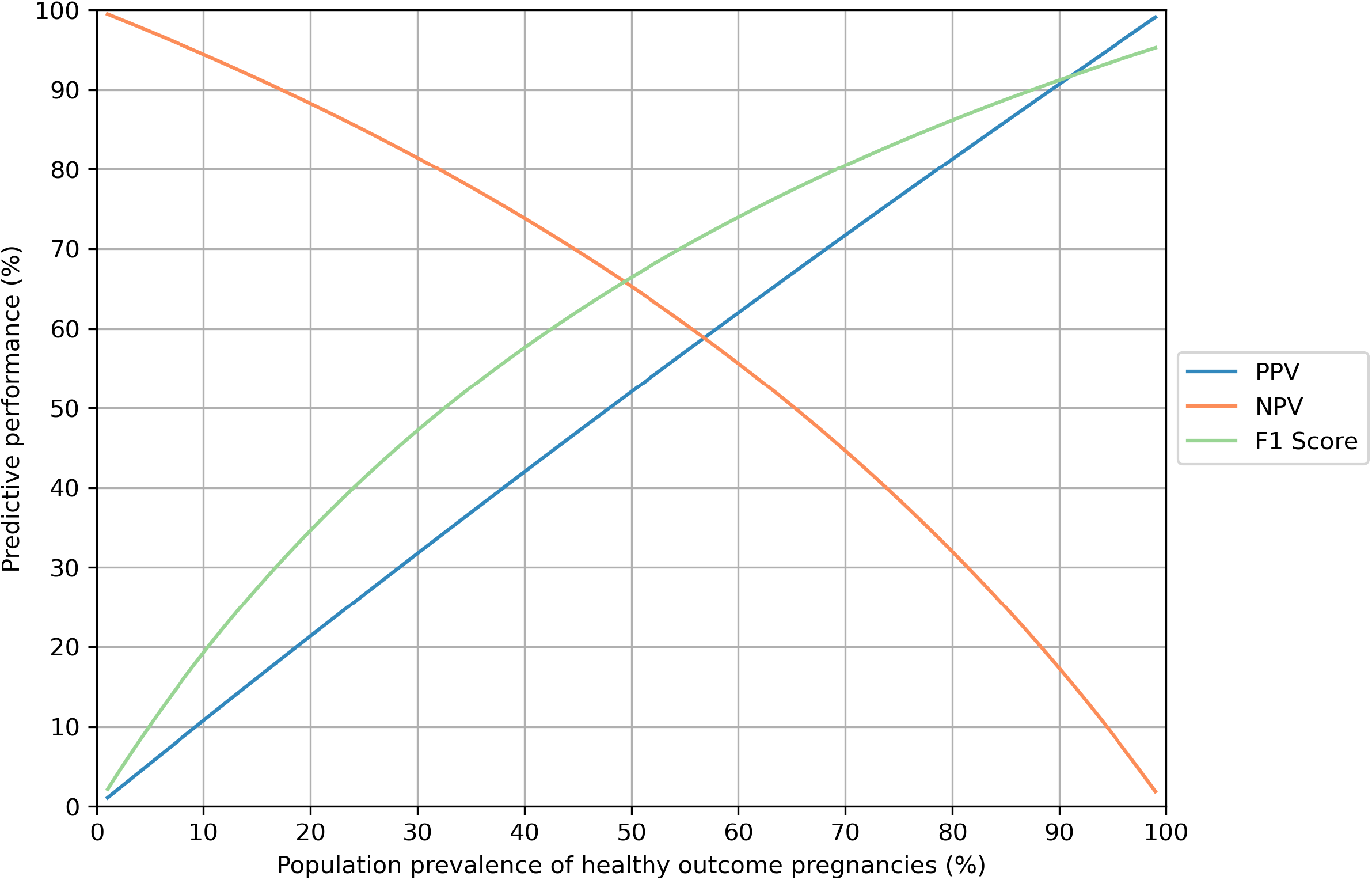
Performance of the Oxford Dawes-Redman system with changing prevalence of healthy outcome pregnancies delivered within 48 hours of FHR monitoring. As the prevalence of healthy outcome pregnancies changes, the positive predictive value (PPV), negative predictive value (NPV) and F1 score change accordingly.

### Temporal proximity to delivery

We then evaluated whether there was a difference in the algorithm’s discriminatory power when comparing FHR traces acquired between 0–24 hours and 24–48 hours prior to delivery (Table 3). The algorithm demonstrated a statistically significant higher accuracy within the 0–24 hour window (56.6%, 95% CI: 55.2–58.1) compared to the 24–48 hour window (51.3%, 95% CI: 50.0–52.6; p=0.008). Sensitivity remained consistently high across both intervals (92.2% for 0–24 hours and 91.9% for 24–48 hours) without significant variation (p=0.898). However, specificity decreased from 21.1% in the 0–24 hour interval to 10.7% in the 24–48 hour interval (p=0.001). Confusion matrices for these time windows are shown in Supplementary Tables 6 and 7.

**Table 3:**
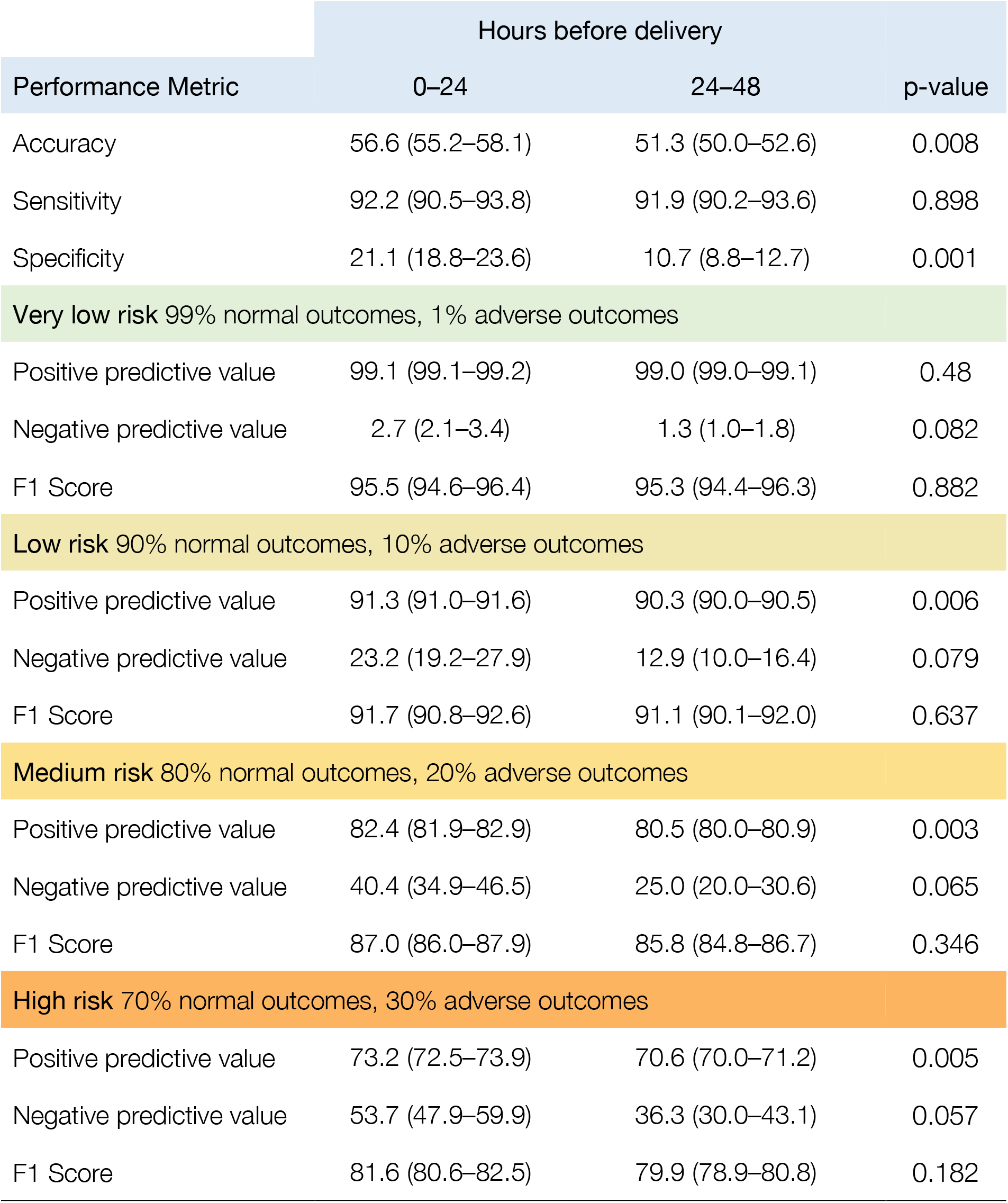
Comparing the Performance of the Oxford Dawes-Redman algorithm at identifying healthy and adverse outcome pregnancies between 0–24 and 24–48 hours before delivery. Analysis was performed for two time windows: those in which a FHR trace was performed between 0–24 and 24–48 hours prior to delivery of the adverse outcome pregnancy. Each adverse outcome FHR trace was matched by gestational age and fetal sex with a corresponding trace from a healthy outcome pregnancy, not necessarily delivered within the same time window. Four theoretical risk groups were evaluated: very low risk, low risk, medium risk and high risk to demonstrate how prevalence of outcomes affects performance. These groups denote the underlying prevalence of a ‘normal’ and ‘adverse outcome’ pregnancy and demonstrate how the performance of the algorithm changes when the prevalence of normality decreases in the population while the prevalence of adverse outcomes increase. As prevalence of normal pregnancies decrease, the algorithm’s ability to identify babies in a state of wellbeing declines substantially.

In the very low-risk group, there was no significant difference between the time windows in terms of sensitivity (p=0.48). A significant decrement in PPV was observed as the risk category increased from low to high, with PPVs of 91.3% and 90.3% in the low-risk category (p=0.006), and from 82.4% to 80.5% in the medium-risk category (p=0.003). In the high-risk category, the PPV declined from 73.2% to 70.6% (p=0.005), and although there was a decrease in the F1 Score from 81.6% to 79.9%, this was not significant (p=0.182).

### Performance by adverse outcome

The Dawes-Redman algorithm’s ability to identify babies in a state of wellbeing did not vary meaningfully when evaluated across the ten discrete outcomes comprising the adverse outcome pregnancy cohort. The algorithm’s accuracy was relatively consistent, across the range of outcomes, with the highest recorded for hypoxic-ischemic encephalopathy (HIE) at 58.0% and the lowest for neonatal sepsis and stillbirth at 50.0% (Supplementary Table 8). The sensitivity for identifying healthy, well babies was high across all conditions with a range of 83.4–99.7%. Specificity remained low across all outcomes, with respiratory conditions at the higher end (22.3%) and stillbirth at the lower end (0.3%).

PPV remained consistently high at 99% or above for all conditions in the low-risk cohort. NPV varied more widely, with a higher value in HIE (28.4%). The F1 Scores were robust across all conditions, reflecting the algorithm’s balanced detection capability wellbeing. These findings confirm the Dawes-Redman algorithm is suitable across a range of conditions when tasked with identifying fetal wellbeing.

## Discussion

We detail here a comprehensive performance evaluation of the Dawes-Redman algorithm in term pregnancies and describe limitations. The Dawes-Redman algorithm is currently integrated into electronic fetal heart rate monitoring systems by principal manufacturers on a global scale.^29,30,51^ in the United Kingdom, this algorithm is utilised in upwards of 120 healthcare institutions and internationally it is distributed to over 110 nations. This system is routinely employed in clinical decision making. However, there has been scant robust analysis of its performance using widely accepted clinical metrics, despite its prominence in obstetric care and national guidelines.^32^

The results described in this study provide crucial insights for obstetric clinical practice. While the accuracy of the Dawes-Redman algorithm (its ability to correctly discern between healthy pregnancies and adverse outcome pregnancies) is relatively low, its sensitivity (ability to correctly identify healthy pregnancies) is high, exceeding 90% on average. When the Dawes-Redman criteria are met, it is a reliable indicator of the fetus being in a state of wellbeing. We have also demonstrated the algorithm is robust in identifying healthy pregnancies across an array of adverse outcomes and when evaluated with increasing time prior to delivery. However, the specificity of the algorithm (ability to correctly identify adverse outcome pregnancies) is low. It is important to note the Dawes-Redman algorithm was not originally developed, and therefore not intended, to identify at-risk pregnancies. Thus, a high degree of caution should be maintained when interpreting any FHR trace failing to meet the Dawes-Redman criteria, and this finding should therefore prompt further assessment of the fetus by other modalities.

The underlying prevalence of healthy pregnancies in the obstetric population is crucial and must be acknowledged in clinical practice. The population prevalence of healthy pregnancies varies throughout clinical practice, dependent on factors such as obstetric history, clinical presentation and examination – even which clinic the pregnant woman is attending. Both positive and negative predictive values (PPV and NPV) are dependent on the prevalence of the outcome of interest – here fetal wellbeing. The PPV in the very low risk population (99% healthy outcomes) was greater than 99%. Therefore, when evaluating the F1 score (the average of sensitivity and PPV), the performance of the system approximated 95%. In these scenarios, the Dawes-Redman algorithm is more reliable. However, we have shown that predictive performance declines when the population prevalence of fetal wellbeing decreases. In a high risk cohort (30% adverse outcome and 70% healthy outcome), the PPV decreased to 73% and consequently the F1 score receded to 80%. In this scenario, the utility of the Dawes-Redman system is more limited. It is important users of this algorithm are aware of this. While it is neither pragmatic nor reasonable for the clinician to know exact prevalences in routine practice, this study demonstrates why appreciating the effect of prevalence is paramount.

We have therefore developed the following conclusions and recommendations:

1. The Dawes-Redman algorithm performs well in its intended purpose: identifying a fetus in a state of wellbeing, i.e. not at immediate risk of an adverse outcome. This is particularly true in a population for which the underlying probability of a normal outcome for the pregnancy is already high – its most common clinical application.
2. Diagnostic performance of any clinical investigation is dependent on the underlying probability (i.e. prevalence) of the outcome of interest. Modelling the effect of prevalence on the performance of the DR algorithm confirms this is just as true for the Dawes-Redman algorithm. The practitioner should consider the pre-test probability of normality on a case-by-case basis when interpreting the results of this system.
3. Failure of a pregnancy to meet the Dawes-Redman criteria is of limited clinical utility in the context of predicting adverse pregnancy outcomes. The ten Dawes-Redman criteria are strict. Failure to meet these criteria is a non-specific indicator of marginally increased risk. Expert clinical opinion always supersedes the Dawes-Redman algorithm. Any doubt or uncertainty surrounding the wellbeing of a fetus should always be an indication to the practitioner to seek expert obstetric opinion, regardless of the result from the Dawes-Redman algorithm.
4. There is a pressing need for the development of FHR analysis algorithms aimed at identifying at-risk or high-risk pregnancies.

This is a robust, large-scale study, however there are important limitations. While the data were acquired over 30 years of obstetric care using myriad fetal heart rate monitoring devices from different manufacturers, a multicentre, ideally international, study would further bolster these results. We also note that the longevity of a Dawes-Redman ‘criteria met’ result is not yet known. Therefore the results of this study should not be extrapolated beyond the 48 hour window we have examined here. This study utilised retrospective data. While the data were collected in a pseudo-prospective manner, i.e. a dedicated team of midwives and clinicians validated and entered these routine clinical data into a dedicated study database on a weekly basis, the gold standard would be a prospective multicentre study. Given the relative paucity of certain adverse obstetric outcomes (e.g. stillbirth occurs in 3.9 births per 1,000 total births in the UK), we estimate the sample size needed for such an undertaking would need approximately eight years. We have already begun such an undertaking at an international obstetric hospital.

## Data Availability

The authors acknowledge the importance of data transparency and the potential value of data sharing in advancing scientific research. However, due to the identifiable and sensitive nature of the data used in this study, which includes detailed fetal heart rate traces potentially linked to individual patient outcomes, we are unable to make the dataset publicly available. The data contains protected health information and is subject to strict confidentiality constraints to safeguard the privacy of individuals. Consequently, the ethical and legal restrictions prevent the sharing of the dataset.

## Acknowledgements

The authors wish to thank Mr Pawel Szafranski and Mr James Bland for their support in providing data acquisition and management services.

