## Supplementary Data for "A Performance Evaluation of Computerised Antepartum Fetal Heart Rate Monitoring: The Dawes-Redman Algorithm at Term"

| | Healthy outcome | Adverse outcome | p-value | Effect size |
|---|---|---|---|---|
| Pregnancies | 1,820 | 1,560 | | |
| Maternal age (years, IQR) | 31.0 (27.0–34.0) | 31.0 (27.0–35.0) | 0.39 | 0.1 (small) |
| Maternal BMI (kg/m$^2$, IQR) | 23.3 (21.2–25.7) | 25.3 (22.2–30.1) | <0.01 | 1.8 (large) |
| Viable parity (IQR) | 1.0 (0.0–1.0) | 0.0 (0.0–1.0) | <0.01 | 0.1 (small) |
| Non-viable parity (IQR) | 0.0 (0.0–1.0) | 0.0 (0.0–1.0) | <0.01 | 0.1 (small) |
| **Smoking status at delivery** (N, %) | | | | |
| Current smoker | 0 (0.0) | 73 (4.7) | <0.001 | 0.2 (medium) |
| Ex-smoker | 335 (18.4) | 142 (9.1) | | |
| Never smoked | 1348 (74.1) | 614 (39.4) | | |
| Non-smoker (history unknown) | 126 (6.9) | 69 (4.4) | | |
| Unknown | 11 (0.6) | 662 (42.4) | | |
| **Labour type** (N, %) | | | | |
| Induction | 613 (33.7) | 800 (51.3) | <0.001 | 0.20 (medium) |
| No labour | 231 (12.7) | 366 (23.5) | | |
| Spontaneous | 976 (53.6) | 394 (25.3) | | |

**Supplementary Table 1: Maternal and labour demographics.** For each fetal monitoring record from an adverse outcome pregnancy, a corresponding healthy control record was identified using propensity score matching, balanced for gestational age at recording and fetal sex. In total, 4,196 FHR monitoring records were identified from each group, developed from 1,820 healthy control pregnancies and 1,560 adverse outcome pregnancies.

| | Normal outcome | Adverse outcome | p-value | Effect size |
|---|---|---|---|---|
| Male (%) | 1,034 (56.8) | 907 (58.1) | 0.46 | 0.01 (small) |
| Female (%) | 786 (43.2) | 653 (41.9) | 0.46 | 0.01 (small) |
| Birthweight (grams, IQR) | 3509.0 (3025.0–3992.0) | 3172.0 (2396.0–3948.0) | <0.001 | 167.7 (large) |
| Gestational age at monitoring ($weeks^{days}$, IQR) | $39^{+1}$ ($38^{+0}$–$40^{+3}$) | $39^{+1}$ ($37^{+6}$–$40^{+4}$) | 0.99 | 0.1 (small) |
| Gestational age at delivery ($weeks^{days}$, IQR) | $40^{+1}$ ($39^{+1}$–$41^{+1}$) | $39^{+3}$ ($38^{+1}$–$40^{+5}$) | <0.001 | 2.2 (large) |
| Apgar scores (IQR) | | | | |
| 1 minute | 10.0 (9.0–10.0) | 9.0 (6.0–10.0) | <0.001 | 0.9 (large) |
| 5 minutes | 10.0 (10.0–10.0) | 10.0 (9.0–10.0) | <0.001 | 0.4 (large) |
| 10 minutes | 10.0 (10.0–10.0) | 10.0 (10.0–10.0) | <0.001 | 0.2 (medium) |
| Delivery method (N, %) | | | | |
| Breech | 0 (0.0) | 4 (0.3) | <0.001 | 0.20 (medium) |
| Forceps | 319 (17.5) | 147 (9.4) | | |
| Emergency caesarean | 0 (0.0) | 529 (33.9) | | |
| Elective caesarean | 231 (12.7) | 366 (23.5) | | |
| Spontaneous vertex | 1184 (65.1) | 450 (28.8) | | |
| Unknown | 0 (0.0) | 1 (0.1) | | |
| Ventouse | 86 (4.7) | 63 (4.0) | | |

Supplementary Table 2: Fetal and delivery demographics.

| Adverse Outcome | Number of affected pregnancies | Number of FHR traces |
|---|---|---|
| Acidaemia | 107 | 182 |
| Asphyxia | 59 | 95 |
| Birthweight <3rd centile | 585 | 969 |
| Extended SCBU admission | 401 | 689 |
| Hypoxic ischaemic encephalopathy | 45 | 79 |
| Low Apgar score | 193 | 360 |
| Neonatal sepsis | 6 | 10 |
| Perinatal infections | 738 | 1188 |
| Respiratory conditions | 675 | 1228 |
| Stillbirth | 12 | 17 |

Supplementary Table 3: Frequency of adverse pregnancy outcomes and associated FHR traces recorded within 48 hours of delivery.

| Adverse Outcome | Definition |
|---|---|
| Acidaemia | Two sets of acidaemia values are used:<br>Babies delivered by CS without labour: arterial pH <7.13 AND arterial BD >10.0<br>Babies who experienced labour (regardless of delivery method): arterial pH <7.05 and arterial BD >14.0. |
| Birth Asphyxia | Low Apgar score(s) (see below) + Acidaemia |
| Birthweight <3$^{rd}$ centile for gestational age | As defined in: Yudkin, P. L., Aboualfa, M., Eyre, J. A., Redman, C. W. G., & Wilkinson, A. R. (1987). New birthweight and head circumference centiles for gestational ages 24 to 42 weeks. Early human development, 15(1), 45-52. |
| Extended SCBU admission | Neonates born at or after $37^{+0}$ gestational weeks who were admitted for at least 7 days to special care baby unit (SCBU) or neonatal intensive care unit (NICU). |
| Hypoxic ischaemic encephalopathy | Diagnosed by treating clinical team (neonatal/paediatrics) |
| Low Apgar score(s) | Apgar score <4 at 1 minute<br>Apgar score <7 at 5 minutes |
| Neonatal sepsis | Diagnosed by treating clinical team (neonatal/paediatrics) |
| Perinatal infection(s) | Phecode X (Extended) NB_856. https://phewascatalog.org/phecode_x. Accessed October 4, 2023. |
| Respiratory conditions | Phecode X (Extended) NB_854. https://phewascatalog.org/phecode_x. Accessed October 4, 2023. |
| Stillbirth | Antepartum or intrapartum stillbirth, as diagnosed by treating clinical team. |

Supplementary Table 4: Definitions of adverse outcomes utilised for inclusion of pregnancy in the adverse outcome cohort.

| | | Pregnancy outcome | |
|---|---|---|---|
| | | Healthy | Adverse |
| Dawes-Redman | Criteria met (positive) | True positive | False positive |
| | Criteria not met (negative) | False negative | True negative |

**Supplementary Table 5: Definitions in a Confusion Matrix.** The Dawes-Redman algorithm categorises fetal heart rate (FHR) traces as 'positive' when criteria indicating fetal wellbeing are met and 'negative' when they are not. In this context, 'positive' corresponds to a healthy pregnancy outcome, while 'negative' indicates otherwise. A 'true positive' is an outcome where the FHR trace is positive and the pregnancy is healthy. A 'false positive' is when the FHR trace is positive but the pregnancy outcome is adverse. A 'true negative' occurs when both the FHR trace and the pregnancy outcome are negative. A 'false negative' is when the pregnancy is healthy but the FHR trace does not meet the criteria. This table serves as a confusion matrix to represent these definitions.

| Dawes-Redman | Pregnancy outcome: Healthy | Pregnancy outcome: Adverse |
|---|---|---|
| Criteria met (positive) | 46.1% (45.2–46.9) | 39.4% (38.2–40.6) |
| Criteria not met (negative) | 3.9% (3.1–4.8) | 10.6% (9.4–11.8) |

**Supplementary Table 6: Confusion Matrix for Dawes-Redman Analysis at term when the Adverse Pregnancy Outcome FHR trace was acquired between 0–24 hours prior to Delivery.** The confusion matrix was developed following bootstrapping. Values are expressed as a percent (%) with 95% confidence intervals.

| | | Pregnancy outcome | |
|---|---|---|---|
| | | Healthy | Adverse |
| Dawes-Redman | Criteria met (positive) | 46.0% (45.1–46.8) | 44.7% (43.7–45.6) |
| | Criteria not met (negative) | 4.0% (3.2–4.9) | 5.3% (4.4–6.3) |

**Supplementary Table 7: Confusion Matrix for Dawes-Redman Analysis at term when the Adverse Pregnancy Outcome FHR trace was acquired between 24–48 hours prior to Delivery.** The confusion matrix was developed following bootstrapping. Values are expressed as a percent (%) with 95% confidence intervals.

| Performance Metric | Acidaemia | Asphyxia | BW<3rd centile | Extended SCBU | HIE | Low Apgar score | Neonatal sepsis | Perinatal infections | Resp. conditions | Stillbirth |
|---|---|---|---|---|---|---|---|---|---|---|
| Accuracy | 51.2 (48.5–53.9) | 51.3 (48.5–54.2) | 52.9 (50.6–55.1) | 54.9 (52.4–57.3) | 58.0 (55.0–61.0) | 52.0 (49.1–54.9) | 50.0 (47.0–53.0) | 51.8 (49.5–54.1) | 56.9 (54.2–59.5) | 50.0 (50.0–50.0) |
| Sensitivity | 91.7 (87.7–95.4) | 94.9 (90.5–98.5) | 92.3 (89.5–94.9) | 91.7 (88.7–94.6) | 98.9 (95.9–99.7) | 87.1 (83.0–91.0) | 83.4 (79.4–86.9) | 90.2 (86.9–93.1) | 91.4 (88.4–94.3) | 99.7 (99.7–99.7) |
| Specificity | 10.6 (7.2–14.7) | 7.7 (3.6–12.3) | 13.6 (10.0–17.2) | 18.0 (14.1–22.1) | 17.1 (12.1–22.4) | 16.8 (12.9–21.1) | 16.6 (12.9–20.6) | 13.3 (10.0–17.0) | 22.3 (18.0–26.5) | 0.3 (0.3–0.3) |
| PPV (99%) | 99.0 (99.0–99.1) | 99.0 (99.0–99.1) | 99.1 (99.0–99.1) | 99.1 (99.1–99.2) | 99.2 (99.1–99.2) | 99.0 (99.0–99.1) | 99.0 (98.9–99.1) | 99.0 (99.0–99.1) | 99.1 (99.1–99.2) | 99.0 (99.0–99.0) |
| NPV (99%) | 1.3 (0.7–2.4) | 1.8 (0.6–5.3) | 1.8 (1.1–2.8) | 2.2 (1.5–3.4) | 28.4 (3.7–46.3) | 1.3 (0.9–2.0) | 1.0 (0.7–1.4) | 1.4 (0.9–2.1) | 2.6 (1.8–3.9) | 1.0 (1.0–1.0) |
| F1 Score (99%) | 95.2 (93.0–97.2) | 96.9 (94.6–98.7) | 95.5 (94.0–96.9) | 95.2 (93.6–96.8) | 99.0 (97.5–99.5) | 92.7 (90.3–94.9) | 90.5 (88.1–92.6) | 94.4 (92.6–96.0) | 95.1 (93.5–96.7) | 99.4 (99.4–99.4) |

**Supplementary Table 8: The performance of the Dawes-Redman algorithm across 10 adverse pregnancy outcomes in FHR traces acquired within 48 hours of delivery.** Each trace from an adverse outcome pregnancy was matched with a trace from a healthy outcome pregnancy using gestational age and fetal sex. The performance metrics remained relatively stable across all outcomes. BW<3rd centile = birthweight <3rd centile for gestational age, Extended SCBU = Extended SCBU admission, HIE = hypoxic ischaemic encephalopathy, Resp. conditions = respiratory conditions, Stillbirth = antepartum or intrapartum stillbirth.
